# Prevalence and associated factors of hypothermia among neonates admitted to the premature baby unit of a secondary care hospital in Sri Lanka: a cross-sectional analytical study

**DOI:** 10.1101/2024.11.15.24317379

**Authors:** Savindi Kasturiarachchi, Inosha Alwis, Sanath Kumara, Buwanaka Rajapaksha

## Abstract

**Background:** Hypothermia is defined as core body temperature being below 36.5°C. This study aimed to identify the prevalence, associated factors and outcomes of hypothermia among neonates admitted to the premature baby unit (PBU) of a secondary care hospital in Nawalapitiya, Sri Lanka.

**Methods:** In a cross-sectional analytical study, medical records of neonates admitted to the PBU were selected retrospectively from March 2022 using consecutive sampling. The recorded axillary temperature on admission, socio-demographic and clinical data were extracted. Following bivariate analysis, multivariable logistic regression was performed.

**Results:** Among 407 neonates, the median admission age was 1 day. The majority were males (52.6%), were term (59%) and had a normal birth weight (52.5%). The prevalence of hypothermia was 38.6% (95% confidence interval (CI):33.9–43.4). Maternal factors like teenage pregnancy, multiple pregnancy, hypertension during pregnancy, premature rupture of membranes and lower-segment caesarean sections; neonatal factors like age on admission being less than 24 hours, prematurity, corrected gestational age on admission being less than 37 weeks, low birth weight, weight on admission being less than 2.5 kg and having been resuscitated at birth had statistically significant associations with hypothermia on the bivariate analysis. Hypothermia showed no significant association with the month of admission. Following multivariable analysis, age on admission being less than 24 hours (adjusted odds ratio (aOR):3.3, 95% CI:1.9-5.8), teenage pregnancy (aOR:8.2, 95% CI:1.8-37.2), multiple pregnancy (aOR:2.8, 95% CI:1.1-7.1) and hypertension in pregnancy (aOR:2.3, 95% CI:1.2-4.7) remained statistically significant. Neonates with hypothermia had 5.2 times (95% CI:1.8-14.6) odds of mortality and 4.9 times (95% CI:2.8-8.5) odds of receiving ventilatory support compared to normothermic neonates. Hypothermia also showed statistically significant associations with infant respiratory distress syndrome, metabolic acidosis and neonatal jaundice.

**Conclusions:** Nearly two out of five neonates admitted to the PBU were hypothermic. There were significant maternal and neonatal associations to be addressed. Hypothermia on admission may indicate serious neonatal morbidity and mortality.

**Summary box:** **What is already known about this topic**

Though hypothermia is known to be associated with neonatal morbidity and mortality, evidence of hypothermia on admission to healthcare settings is limited, especially in South Asia.

**What this study adds**

This study adds evidence on the prevalence of admission hypothermia among newborns, and the maternal, neonatal and environmental factors independently associated with it. It also highlights the specific neonatal complications to which hypothermia is linked.

**How this study might affect research, practice or policy**

These findings can be used to identify risk groups during neonatal care and inform advocacy and policy for thermo-protective interventions within hospitals.

## Introduction

Hypothermia is defined as a core body temperature that is less than 36.5 degrees centigrade. In adult humans, hypothermia develops due to prolonged exposure to cold temperatures and leads to multiple systemic complications. Neonates – newborns who are less than 29 days of age – are at a higher risk for hypothermia than adults. Hypothermia manifests in neonates even without major exposure to cold temperatures and leads to greater morbidity and mortality. It is estimated that annually 17 million neonates become hypothermic in low- and middle-income (LMIC) countries, mostly in countries with a higher burden of neonatal mortality (1). Neonatal hypothermia is not limited to countries with cold climates but is similarly prevalent in tropical regions (2–5). It has also been identified as a neglected challenge worldwide (6). These facts underscore the importance of addressing neonatal hypothermia in the global agenda for newborn care.

The vulnerability of neonates to hypothermia is related to physiological factors in the neonatal period. In utero, the foetus can produce heat as a byproduct of cellular respiration at a rate of 33-47 Calories/kg/min. After birth, heat loss mechanisms including radiation, evaporation, convection and conduction take effect. The rate of heat loss may be as high as 100-200 Calories/kg/min resulting in a temperature drop of 0.2-1℃/min (7,8). Though term neonates can generate heat up to two times the foetal rate, it is still inadequate to maintain normothermia, particularly on the first day of life (8).

Neonates are also at a higher risk of hypothermia as they have a large surface area compared to body mass. The surface area of a neonate is three times that of an adult. This results in a greater heat loss in comparison to heat generation. The skin of a neonate is thin and heat permeable, causing increased water loss in the first few days of life. There is also very little subcutaneous fat resulting in poor heat insulation. The neonate has poorly developed autonomic and chemical pathways including poor heat conservation mechanisms such as shivering or curling up in response to cold environments (9–11).

As these risk factors are more prominent among preterm neonates, they are at an even higher risk of developing hypothermia in their early days of life. Hypothermia can further occur during routine baby care following delivery, during resuscitation or when transporting the baby to the baby unit. Therefore, studying the prevalence and associated factors of hypothermia among neonates who are admitted to neonatal care units, commonly referred to as ‘admission hypothermia’, has been given an important focus in the literature.

Most studies in the literature about admission hypothermia have been conducted in the African region. A meta-analysis by Beletew et al. (2020) showed a pooled prevalence of 57.2% of neonatal hypothermia among hospital-based studies in East Africa (5). Phoya et al. (2020) detected 74% of neonates in a tertiary hospital in Malawi to be hypothermic 5 min after birth while 77% were found to be hypothermic on admission to the neonatal unit (12). Evidence on the burden of neonatal hypothermia in healthcare settings is scarce in South Asia as most studies in the region have targeted community settings (6). Among limited available studies, a study in Nepal showed a hypothermia prevalence of 64% among babies in a newborn unit (13). Only one study in Sri Lanka had assessed the prevalence of hypothermia among admitted neonates previously. Madhvi et al. (2014) conducted a study involving two hospitals in Western Sri Lanka which reported a prevalence of 63% of admission hypothermia (14).

There are neonatal, maternal, institutional and environmental risk factors for neonatal hypothermia. The systematic review by Beletew et al. (2020) identified low birth weight, prematurity and medical problems as neonatal risk factors; caesarean section delivery, obstetric complications during pregnancy and labour were observed as maternal risk factors (5). Several studies in African countries reported early bathing, absence of immediate skin-to-skin contact with the mother after birth, delay in initiation of breastfeeding, night-time delivery and admissions during the cold season as common institutional and environmental risk factors for hypothermia on admission to neonatal intensive care units (2,3,15–21).

Though hypothermia is rarely identified as a direct cause of death among newborns, it contributes significantly towards neonatal morbidity and mortality by acting as a comorbidity (6). It is associated with the most common causes of neonatal deaths worldwide: neonatal infections, prematurity and birth asphyxia (6). Neonatal hypothermia leads to increased energy consumption, hypoxia, reduced peripheral circulation causing tissue ischaemia, renal failure, necrotising enterocolitis, coagulation defects and sepsis (22). Nayeri et al. (2006) reported a significant association between hypothermia at birth and respiratory distress in the first six hours of life, jaundice, hypoglycaemia and metabolic acidosis among newborns in Iran, regardless of birth weight and gestational age (22).

Considering mortality, a study conducted in a health facility in Nepal showed an 80% rise in mortality for every 1-degree Celsius decrease in neonatal body temperature (23). Johanson et al. (1993) reported that 15.6% of neonates who were hypothermic on admission to a neonatal unit in Kathmandu, Nepal died indicating a significant association between hypothermia on admission and mortality (13). Neonates with hypothermia had 2 to 3 times higher mortality rates than normothermic babies on admission to neonatal intensive care units (3,4).

Given the serious morbidity and mortality associated with neonatal hypothermia, generating epidemiological evidence on its prevalence, associated factors and consequences will be imperative in designing health interventions for early detection and prevention, especially in low-resource settings. As stated before, there is also a critical gap in knowledge concerning the burden and associations of admission hypothermia in Sri Lankan and South Asian healthcare settings. Our findings will thus support policy actions and advocacy for newborn care in the region. Therefore, the overall aim of this study was to determine the prevalence, associated factors and outcomes of hypothermia among neonates admitted to the PBU of a secondary care hospital in Sri Lanka.

## Methods

### Study design and setting

This was a cross-sectional analytical study conducted at the PBU of the District General Hospital (DGH) Nawalapitiya. DGH Nawalapitiya is a secondary care hospital that is located in the Kandy district of Central Province, Sri Lanka. There are about 200 deliveries that take place at DGH Nawalapitiya per month and approximately 40 neonates are admitted to the PBU monthly. It must be noted that contrary to the name of the unit, its care is not limited to premature babies. PBU of DGH Nawalapitiya provides services to any neonate – either term or preterm – who requires specialised neonatal care. It consists of facilities standard to any neonatal care unit in Sri Lanka including incubators, warmers and ventilators. This study was conducted at the PBU from April 2022 to November 2023. Data collection was carried out retrospectively starting from March 2022. Ethical clearance was obtained from the Ethical Review Committee of the National Hospital Kandy (ERC No: NHK/ERC/19/2022). Necessary administrative permissions were taken from DGH Nawalapitiya. This study did not involve codesigning with patients or the public.

### Study population and sampling

All neonates (infants who are aged 1 to 28 days) admitted to the PBU of DGH Nawalapitiya with a measurement of axillary temperature using a digital thermometer were included in the study. Readmissions of the same neonate were excluded.

The minimum sample size was calculated using the formula to estimate a population proportion with specified absolute precision (24). For a 95% confidence level, 5% precision and an expected proportion of 0.5 to estimate the largest sample size for an unknown proportion of hypothermia among neonates admitted to the PBU, the minimum sample size was computed as 384. This was inflated by 10% to account for possible missing information in bed head tickets (BHT) related to variables other than the recorded temperature, rendering a sample size of 422. A consecutive sampling technique was followed starting with the last patient admitted on March 2022 and proceeding retrospectively till the required sample size was achieved.

### Study instruments and data collection

The data source was the BHT pertaining to the neonate admitted to the PBU. A structured data collection sheet was used to collect data on patient demographic data, clinical information and measurements of axillary temperature available on the BHT. At the PBU of DGH Nawalapitiya, all axillary temperature measurements are recorded by the admitting nursing officer using a digital thermometer and standard techniques.

A pretest was carried out using a few sample BHTs to ensure the comprehensibility of the structured data collection sheet and to identify other possible issues that may be encountered during the process of data extraction and entering. Neonates fulfilling the inclusion criteria were recruited and the structured data collection sheet was used to extract data from their BHTs. Data collection was performed by a research assistant who was given prior training in secondary data collection using medical records. Extracted data was cross-examined by a co-investigator for accuracy and reporting errors. Ambiguity in data that arose due to the style and legibility of writing in BHTs was resolved through a consensus among co-investigators, or in severe cases, was classified as missing data.

### Data analysis

Data was entered into Microsoft Office Excel and was checked for duplicates and missing records. Complete case analysis was carried out. After the coding of data, the dataset was added to SPSS 25 (Statistical Package for Social Sciences 25.0) for analysis. Operationalisation of hypothermia (outcome variable) was performed according to the World Health Organisation’s definitions and is described in Figure 1 (10). Operationalisation of all neonatal and maternal exposure variables including birth weight and prematurity categories followed standard definitions in paediatric practice and are defined in Table 1 and 2 (9,11).

**Figure 1:**
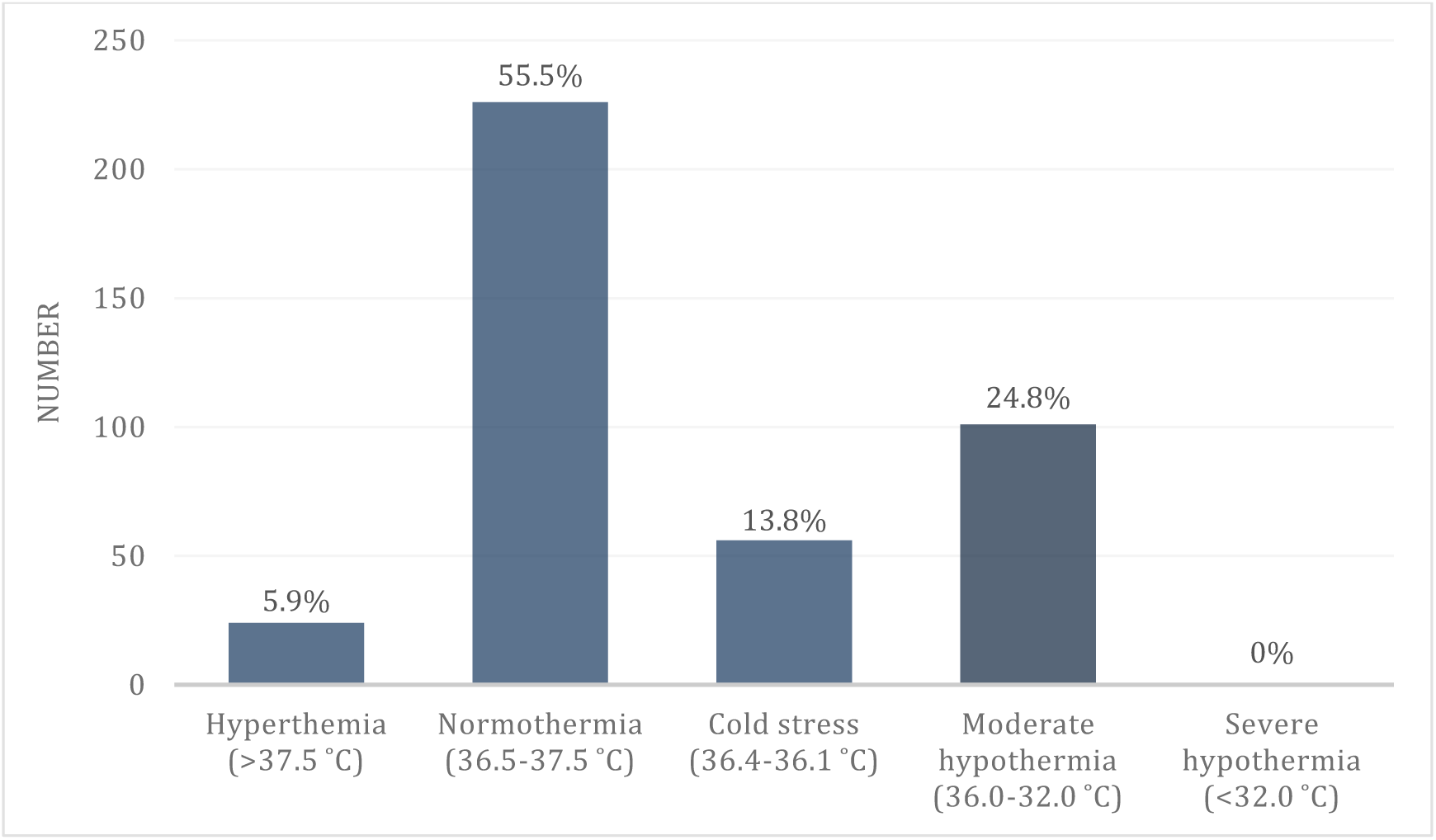
Grades of hypothermia among neonates admitted to the PBU

**Table 1:** Neonatal characteristics.

| **Variable** | **Categories** | **Frequency** | **Percentage** |
| --- | --- | --- | --- |
| **Sex** | Male | 214 | 52.6% |
|  | Female | 192 | 47.2% |
|  | Ambiguous | 1 | 0.2% |
| **Age on admission** | ≤24 hours | 235 | 57.7% |
|  | >24 hours | 172 | 42.3% |
| **Gestational age** | Post term (≥42 weeks) | 0 | 0% |
|  | Term (37 – 41+6 weeks) | 240 | 59.3% |
|  | Late preterm (34 – 36+6) | 95 | 23.5% |
|  | Moderate preterm (32 – 33+6) | 27 | 6.7% |
|  | Very preterm (28 – 31+6) | 37 | 9.0% |
|  | Extremely preterm (<28 weeks) | 6 | 1.5% |
| **Corrected gestational age on admission** | ≥42 weeks | 0 | 0% |
|  | 37 – 41+6 weeks | 259 | 64% |
|  | 34 – 36+6 | 81 | 20% |
|  | 32 – 33+6 | 28 | 6.9% |
|  | 28 – 31+6 | 32 | 7.9% |
|  | <28 weeks | 5 | 1.2% |
| **Birth weight** | Overweight (>3.5 kg) | 18 | 4.4% |
|  | Normal birth weight (3.5 - 2.5 kg) | 213 | 52.5% |
|  | Low birth weight (2.499 – 1.5 kg) | 119 | 29.3% |
|  | Very low birth weight (1.499 – 1 kg) | 43 | 10.6% |
|  | Extremely low birth weight (<1 kg) | 13 | 3.2% |
| **Weight on admission** | >3.5 kg | 21 | 5.2% |
|  | 3.5 - 2.5 kg | 185 | 45.6% |
|  | 2.499 – 1.5 kg | 144 | 35.5% |
|  | 1.499 – 1 kg | 44 | 10.8% |
|  | <1 kg | 12 | 3.0% |
| **Resuscitated at birth** | Yes | 59 | 14.5% |
|  | No | 347 | 85.5% |
| **Admitted from** | Operation theatre | 130 | 31.9% |
|  | Outpatient department | 98 | 24.1% |
|  | Postnatal ward | 97 | 23.8% |
|  | Labour room | 66 | 16.2% |
|  | Transferred from other hospitals | 10 | 2.5% |
|  | Emergency treatment unit | 4 | 1% |
|  | COVID isolation room | 2 | 0.5% |
| **IRDS** | No IRDS | 321 | 79.3% |
|  | Mild IRDS | 72 | 17.8% |
|  | Moderate IRDS | 6 | 1.5% |
|  | Severe IRDS | 6 | 1.5% |
| **Hypoglycaemia** | No hypoglycaemia | 366 | 90.4% |
|  | Hypoglycaemia | 39 | 9.6% |
| **Metabolic acidosis** | No metabolic acidosis | 370 | 91.4% |
|  | Metabolic acidosis | 35 | 8.6% |
| **Neonatal jaundice** | No neonatal jaundice | 260 | 64.2% |
|  | Neonatal jaundice | 145 | 35.8% |
| **Pulmonary haemorrhage** | No pulmonary haemorrhage | 403 | 99.8% |
|  | Pulmonary haemorrhage | 1 | 0.2% |
| **Sepsis** | No sepsis | 353 | 87.2% |
|  | Sepsis | 39 | 9.6% |
|  | Meningitis | 12 | 3.2% |
| **Ventilatory support** | No ventilatory support required | 331 | 82% |
|  | Continuous positive airway pressure | 43 | 10.6% |
|  | Mechanical ventilation | 30 | 7.4% |
| **Outcome** | Alive and discharged | 383 | 94.3% |
|  | Diseased | 20 | 4.9% |
|  | Alive and transferred | 3 | 0.8% |

Descriptive statistics were reported using both numerical and graphical methods. Bivariate analysis of categorical variables was performed using chi square tests. Categorical variables with multiple groups were recoded into dichotomous variables when required. Relevant crude odds ratios (OR) were calculated with their 95% confidence intervals (CI). A probability value (*p* value) of less than 0.05 was considered to be statistically significant.

Multivariable logistic regression was carried out in line with the standards developed by Bagley et al. (2001) for using and reporting logistic regression in the medical literature (25). Only associations that were found to be significant in bivariate analysis were included in the regression model. Each predictor variable was coded in binary groups similar to the bivariate analysis. The number of events per predictor variable was assessed. Multicollinearity between the predictors was evaluated by running collinearity diagnostics using multiple linear regression with the outcome of hypothermia taken as a continuous variable. Variance inflation factor (VIF) over 5 was considered as the cut-off. Since we aimed to develop an explanatory model for neonatal hypothermia in hospital settings rather than a predictive model, all the eligible predictors were fitted simultaneously into the model, without following automated stepwise procedures. The overall model significance and the goodness of fit were determined using the Omnibus test and Hosmer and Lemeshow tests, respectively. Classification accuracy of the model and pseudo R square values were reported. Adjusted odds ratios (aOR) were calculated with their 95% CI. Testing for interactions between the predictors was not performed. The STROBE cross sectional checklist was used when writing our report (26).

## Results

### Study participants

Out of 434 records, 27 were excluded due to missing data (6.2%) and 407 neonates were included in the final analysis. Table 1 shows the characteristics of the study population. The majority (52.6%) were males. One baby was categorised as having ambiguous genitalia. Most neonates (57.7%) were admitted within the first 24 hours of life. The median admission age was 1 day (interquartile range (IQR):4). The majority (59%) were term babies. Among the preterm, more than half (57.3%) were late preterm. 37% of the neonates had a corrected gestational age that was less than 37 on admission. Most neonates (52.5%) had a normal birth weight. The mean birth weight was 2.46 kg (standard deviation (SD): 0.76). 49.3% of the neonates weighed less than 2.5 kg on admission. One out of seven of the babies (14.5%) were resuscitated at birth. The commonest places from where the neonates were admitted were the operation theatre (31.9%), the postnatal ward (23.8%) and the outpatient department (24.1%).

The main morbidities experienced by neonates during admission are also summarised in Table 1. 18% of the sample required ventilatory support. There were 20 deaths (4.9%) reported during the period of admission.

### Characteristics of the mothers

The mean maternal age was 29 years (SD:5.3). 3.2% were teenage mothers (less than 20 years). The median parity of mothers was 2 pregnancies (IQR:2). Over three-fourths (90.1%) had singleton pregnancies. Nearly half (44.9%) were primi mothers. Table 2 summarises the main morbidities experienced by mothers during their pregnancies. The majority (58.2%) were delivered via lower segment caesarean section (LSCS).

**Table 2:**
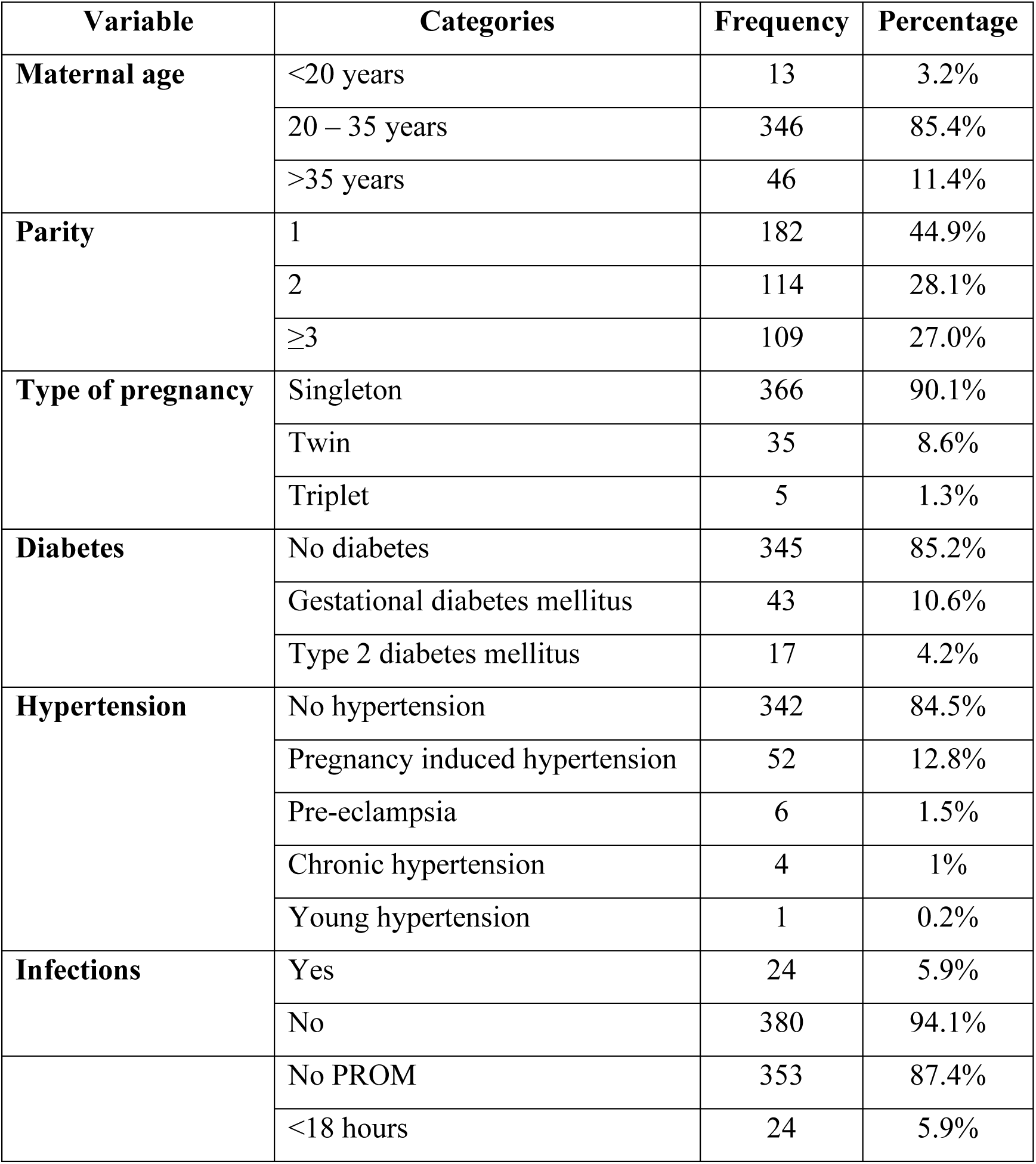
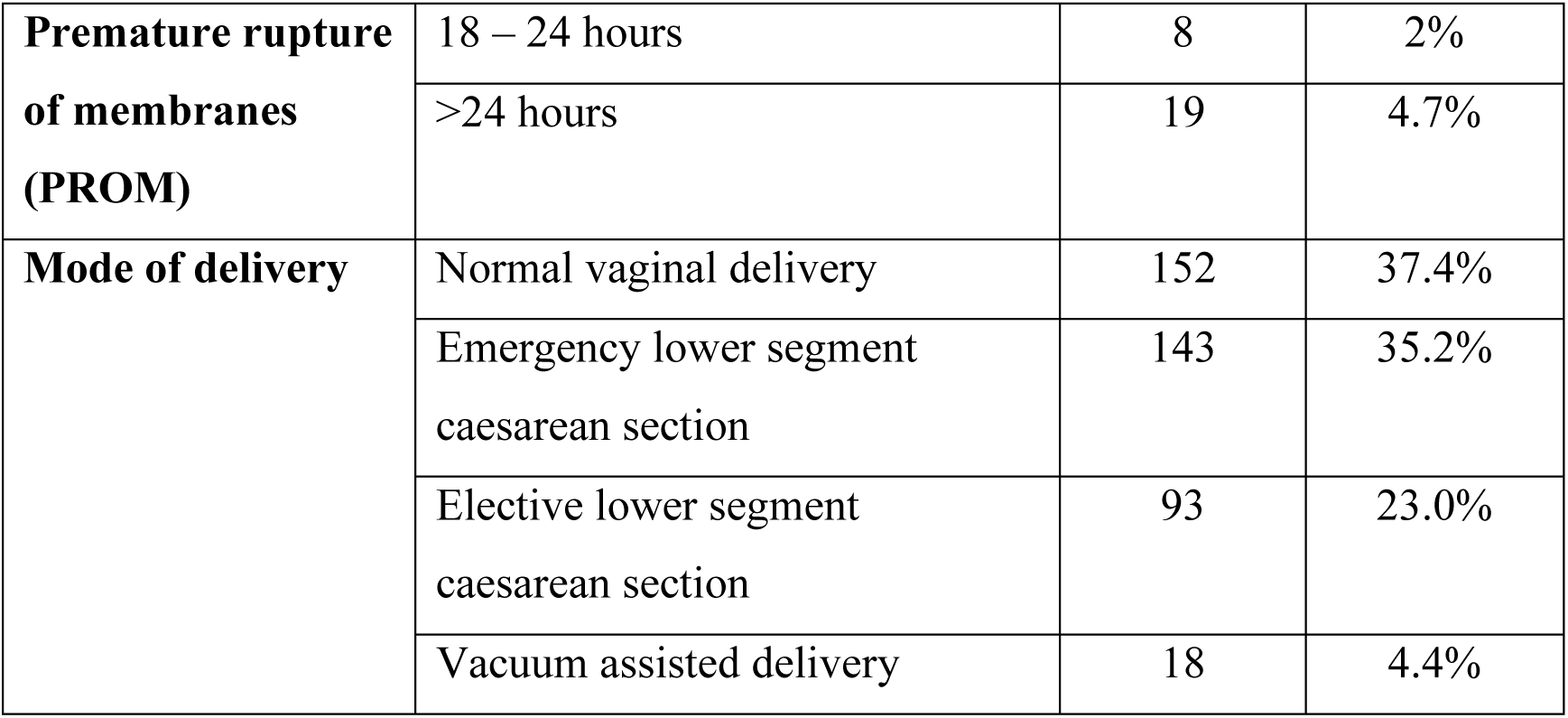
Maternal characteristics.

### Prevalence of neonatal hypothermia

The prevalence of hypothermia among neonates on admission to the PBU was 38.6% (95% CI:33.9–43.4). As seen in Figure 1, of the 157 neonates that were found to be hypothermic, the majority had moderate hypothermia (n=101). There were no severely hypothermic babies.

### Associations of hypothermia

#### Neonatal and maternal factors

As shown in Table 3, in the bivariate analysis, neonatal factors that were significantly associated with admission hypothermia were, admission age being less than 24 hours, prematurity, corrected gestational age on admission being less than 37 weeks, low birth weight, admission weight being less than 2.5 kg and being resuscitated at birth. The largest measure of association with admission hypothermia was observed among neonates whose corrected gestational age was less than 37 weeks (OR: 8.7, 95% CI: 5.5-13.7). Being a female baby was identified as a protective factor in the sample (OR: 0.8, 95% CI: 0.8-1.3) although it was not statistically significant.

**Table 3:** Bivariate and multivariable analysis of associated factors of hypothermia.

| Variable | Hypothermic<br>(157) | Non-<br>hypothermic | OR<br>(95%CI) | $p$<br>value | aOR<br>(95%CI) | $p$<br>value |
| --- | --- | --- | --- | --- | --- | --- |
|  |  | (250) |  |  |  |  |
|  | n (%) | n (%) |  |  |  |  |
| **Neonatal factors** |  |  |  |  |  |  |
| Sex of neonate |  |  |  |  |  |  |
| Female | 77 (40.1%) | 115 (59.9%) | 0.8 | 0.600 |  |  |
| Not female | 80 (37.6%) | 134 (62.4%) | (0.6-1.3) |  |  |  |
| Age on admission (hours) |  |  |  |  |  |  |
| ≤ 24 | 129 (54.9%) | 106 (45.1%) | 6.3 | <0.001 | 3.3 | <0.0 |
| > 24 | 28 (16.3%) | 144 (83.7%) | (3.8-10.1) |  | (1.9-5.8) | 01 |
| Gestational age (weeks) |  |  |  |  |  |  |
| < 37 | 106 (63.5%) | 61 (36.5%) | 6.4 | <0.001 | 1.5 | 0.234 |
| ≥ 37 | 51 (21.2%) | 189 (78.8%) | (4.1-10.0) |  | (0.8-2.9) |  |
| Corrected gestational age on admission (weeks) |  |  |  |  |  |  |
| < 37 | 103 (69.6%) | 45 (30.4%) | 8.7 | <0.001 |  |  |
| ≥ 37 | 54 (20.8%) | 205 (79.2%) | (5.5-13.7) |  |  |  |
| Birth weight (kg) |  |  |  |  |  |  |
| < 2.5 | 110 (62.9%) | 65 (37.1%) | 6.6 | <0.001 | 3.2 | 0.058 |
| ≥ 2.5 | 47 (20.3%) | 184 (79.7%) | (4.3-10.3) |  | (0.9-11.0) |  |
| Weight on admission (kg) |  |  |  |  |  |  |
| < 2.5 | 112 (56%) | 88 (44%) | 4.5 | <0.001 | 0.8 | 0.724 |
| ≥ 2.5 | 45 (21.8%) | 161 (78.2%) | (2.9-7.0) |  | (0.2-2.6) |  |
| Resuscitated at birth |  |  |  |  |  |  |
| Yes | 37 (62.7%) | 22 (37.3%) | 3.2 | <0.001 | 1.8 | 0.104 |
| No | 120 (34.6%) | 227 (65.4%) | (1.8-5.6) |  | (0.9-3.4) |  |
| **Maternal factors** |  |  |  |  |  |  |
| Teenage pregnancy |  |  |  |  |  |  |
| Yes | 10 (76.9%) | 3 (23.1%) | 5.5 | 0.004 | 8.2 | 0.006 |
| No | 147 (37.5%) | 245 (62.5%) | (1.5-20.5) |  | (1.8-37.2) |  |
| Parity |  |  |  |  |  |  |
| Primi | 74 (40.7%) | 108 (59.3%) | 1.2 | 0.437 |  |  |
| Not<br>primi | 83 (36.9%) | 142 (63.1%) | (0.7-1.7) |  |  |  |
| Multiple pregnancy |  |  |  |  |  |  |
| Yes | 31 (77.5%) | 9 (22.5%) | 6.5 | <0.001 | 2.8 | 0.025 |
| No | 126 (34.4%) | 240 (65.6%) | (3.0-14.2) |  | (1.1-7.1) |  |
| Diabetes in pregnancy |  |  |  |  |  |  |
| Yes | 23 (38.3%) | 37 (61.7%) | 0.9 | 0.941 |  |  |
| No | 134 (38.8%) | 211 (61.2%) | (0.5-1.7) |  |  |  |
| Hypertension in pregnancy |  |  |  |  |  |  |
| Yes | 40 (63.5%) | 23 (36.5%) | 3.3 | <0.001 | 2.3 | 0.016 |
| No | 117 (34.2%) | 225 (65.8%) | (1.9-5.8) |  | (1.2-4.7) |  |
| Maternal infections |  |  |  |  |  |  |
| Yes | 11 (45.8%) | 13 (54.2%) | 1.36 | 0.47 |  |  |
| No | 146 (38.4%) | 234 (61.6%) | (0.59-3.1) |  |  |  |
| Premature rupture of membranes |  |  |  |  |  |  |
| Yes | 32 (62.7%) | 19 (37.3%) | 3.0 | <0.001 | 1.5 | 0.288 |
| No | 125 (35.4%) | 228 (64.6%) | (1.6-5.6) |  | (0.7-3.4) |  |
| Mode of delivery |  |  |  |  |  |  |
| LSCS | 101 (42.8%) | 135 (57.2%) | 1.5 | 0.044 | 1.1 | 0.796 |
| Not<br>LSCS | 56 (32.9%) | 114 (67.1%) | (1.0-2.3) |  | (0.6-1.8) |  |

Considering maternal factors, teenage pregnancy, multiple pregnancy, hypertension during pregnancy, premature rupture of membranes and undergoing LSCS were found to have statistically significant associations with neonatal hypothermia in the bivariate analysis as seen in Table 3. Associations with being a primi mother, having diabetes or infections during pregnancy were not statistically significant.

### Environmental factors

The month of admission was the only external environmental factor that was assessed in this study. The highest mean temperature on admission was observed during April (36.57 °C) and the lowest mean temperature was in January (36.07 °C). Figure 2. illustrates the monthly changes in the percentage of neonates with hypothermia. Admission hypothermia was highest during January when over half of the neonates (52.5%) were found to be hypothermic. It was the lowest during April (25.7%). Bivariate analysis showed no significant association with the month of admission (chi square: 14.3, *p* value: 0.215).

**Figure 2:**
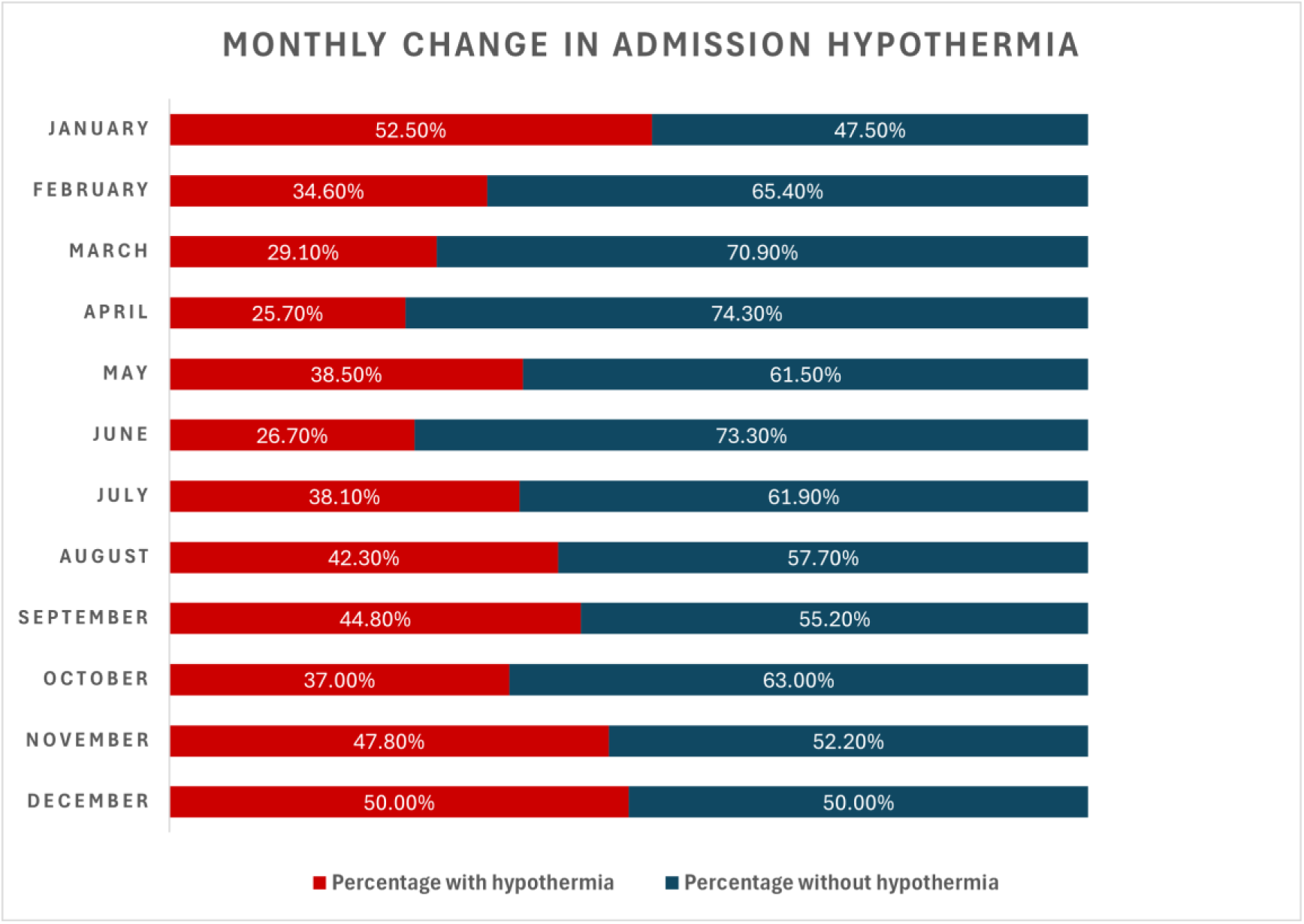
Monthly change in the percentage of neonates with admission hypothermia

### Multivariable regression model

In the multivariable logistic regression model, the number of events per predictor variable was equal to the recommended 10:1 ratio. In collinearity diagnostics, prematurity and corrected gestational age on admission variables were seen to have VIF over 5. Therefore, out of the two, only prematurity was selected for the final model. The model showed satisfactory statistical significance (Omnibus test, chi square: 140.1, *p* value: <0.001) and goodness of fit (Hosmer and Lemeshow test, chi square: 13.64, *p* value:0.09). It had an overall classification accuracy of 76.5% and Nagelkerke pseudo R was 39.8%. In the multivariable analysis, as shown in Table 3, only admission age being less than 24 hours remained statistically significant out of the neonatal factors (aOR:3.3, 95% CI: 1.9-5.8). Among maternal factors, only teenage pregnancy, multiple pregnancy and hypertension in pregnancy maintained statistically significant associations with neonatal hypothermia in the final model with all the predictors. The maternal factor with the largest measure of effect with hypothermia was teenage pregnancy (aOR: 8.2, 95% CI: 1.8-37.2).

### Morbidity and mortality associated with neonatal hypothermia

On analysis of the complications associated with admission hypothermia shown in Table 4, infant respiratory distress syndrome, metabolic acidosis, neonatal jaundice and requiring ventilatory support were found to be statistically significant. Neonates with hypothermia had 4.9 times (95% CI:2.8-8.5) odds of receiving ventilatory support and 5.2 times (95% CI:1.8-14.6) odds of mortality compared to normothermic neonates.

**Table 4:** Analysis of neonatal complications associated with hypothermia.

| Variable | Hypothermic | Non-<br>hypothermic | OR (95%CI) | p value |
| --- | --- | --- | --- | --- |
|  | n (%) | n (%) |  |  |

| Infant respiratory distress syndrome |  |  |  |  |
| --- | --- | --- | --- | --- |
| Yes | 54 (34.4%) | 30 (12.1%) | 3.8 (2.3-6.3) | <0.001 |
| No | 103 (65.6%) | 218 (87.9%) |  |  |
| Hypoglycaemia |  |  |  |  |
| Yes | 19 (12.2%) | 20 (8%) | 1.5 (0.8-3.1) | 0.169 |
| No | 137 (87.8%) | 229 (92%) |  |  |
| Metabolic acidosis |  |  |  |  |
| Yes | 20 (12.8%) | 15 (6%) | 2.3 (1.1-4.6) | 0.018 |
| No | 136 (87.2%) | 234 (94%) |  |  |
| Neonatal jaundice |  |  |  |  |
| Yes | 77 (49.4%) | 68 (27.3%) | 2.6 (1.7-3.9) | <0.001 |
| No | 79 (50.6%) | 181 (72.7%) |  |  |
| Pulmonary haemorrhage |  |  |  |  |
| Yes | 1 (0.6%) | 0 (0%) | 1.0 (0.9-1.0) | 0.204 |
| No | 154 (99.4%) | 249 (100%) |  |  |
| Neonatal sepsis |  |  |  |  |
| Yes | 20 (12.8%) | 31 (12.5%) | 1.0 (0.5-1.8) | 0.925 |
| No | 136 (87.2%) | 217 (87.5%) |  |  |
| Ventilatory support |  |  |  |  |
| Yes | 51 (32.5%) | 22 (8.9%) | 4.9 (2.8-8.5) | <0.001 |
| No | 106 (67.5%) | 225 (91.1%) |  |  |
| Outcome |  |  |  |  |
| Deceased | 15 (9.6%) | 5 (2%) | 5.2 (1.8-14.6) | 0.001 |
| Not deceased | 141 (90.4%) | 245 (98%) |  |  |

## Discussion

This study assessed the prevalence, factors and outcomes associated with neonatal hypothermia in a secondary care setting in Sri Lanka. Our findings showed that nearly two out of every five neonates had admission hypothermia. In the multiple logistic regression model, admission age being less than 24 hours was the only neonatal factor associated with hypothermia; teenage pregnancy, multiple pregnancy and hypertension in pregnancy were the identified maternal factors associated. Infant respiratory distress syndrome, metabolic acidosis, neonatal jaundice and the requirement of ventilatory support were the complications associated with neonatal hypothermia. Hypothermic neonates were five times more likely to die compared to normothermic neonates.

The strengths of our study included calculating the prevalence of admission hypothermia in a large sample of neonates using secondary data and analysing the associated factors and outcomes using a multivariable analysis, thereby minimising the confounding effects. There are several limitations of our study. First, since the study was based on secondary data its internal validity may be subjected to the errors in medical record-keeping process. However, we attempted to minimise that information bias by using a single data extractor, using explicit operationalisation and standard review methods. Second, we were unable to verify whether the nursing officers adhered to the same techniques in measuring temperature or the inter-instrument reliability of the digital thermometers. Third, during multivariable analysis, our model did not account for possible interactions between the predictor variables. Fourth, the cross-sectional nature of this study can only establish associations and not causal risk factors of neonatal hypothermia. Fifth, since this study was carried out in a single centre in the Central Province of Sri Lanka, the generalisability of the prevalence findings to other regions may be limited.

The prevalence of neonatal hypothermia seen in our study was lower than the prevalence seen in healthcare settings around the world. It was lower than the pooled prevalence of hypothermia (57.2%) as reported by a hospital-based systematic review done in Africa and also the 64% prevalence observed in a healthcare facility in Nepal, where the rectal temperature was measured on admission (5,13). Our findings were also lower than previously observed prevalence rates in Sri Lanka (63%) as reported by Madhvi et al. (14). This disparity in prevalence could be due to varying availability of resources in individual healthcare facilities and geographical differences. East African studies have identified possible causes as a lack of appropriate towels and other laundry items for the thermal control of neonates. The lower-than-expected rates of hypothermia in our study may be attributed to protective factors identified in the literature such as the availability of warm towels, plastic bags for delivery of preterm neonates, radiant warmers in the delivery rooms and transport incubators at the PBU (3,13,27). Measurement of rectal temperature in the previous Sri Lankan study could have also given falsely high values compared to the axillary temperature measurement that is recommended by guidelines (28). Nevertheless, more investigations are needed into regional differences in neonatal hypothermia in Sri Lanka.

The only statistically significant neonatal factor identified was the admission age being less than 24 hours, which was similar to the findings of several studies done in hospitals of East and West Africa which showed that such neonates had up to 3.6 times higher odds of developing admission hypothermia (2–4). This could be due to poorly developed thermoregulatory mechanisms in neonates which is even more pronounced in the early hours of life and lack of adipose tissue, thus putting newborns at a higher risk of hypothermia. In the analysis of maternal factors, our findings showed that teenage pregnancy was significantly associated with neonatal hypothermia. A study done in a health facility in Uganda also found that newborns of adolescent mothers were at a significantly higher risk of hypothermia while another study done in a hospital in Nigeria found that maternal age being more than 20 years was a protective factor against neonatal hypothermia on admission (3,21). This may be due to vulnerabilities in the bio-psycho-social environment surrounding teenage pregnancies that challenge the effective provision of warmth to neonates. The second maternal factor found to be significant was multiple pregnancy. This was consistent with a study done in a healthcare setting in Iran which showed 1.6 times the odds of neonatal hypothermia in multiple pregnancy compared to singletons (29). This could be due to inadequate facilities and health staff in the delivery rooms to effectively maintain the warm chain for multiple babies delivered at once. The third significant maternal factor was hypertension during the current pregnancy. A multivariable analytical study done in a hospital in Canada and a study done in a neonatal intensive care unit in Brazil found that neonates of mothers with hypertension during pregnancy and preeclampsia, which is an entity of hypertension in pregnancy, had a significantly higher chance of developing hypothermia (30,31). Three similar hospital-based studies done in Ethiopia have found maternal obstetric complications to be significantly associated with neonatal hypothermia on admission (15,16,32). This could be attributed to the mothers being too ill to adequately care for a newborn and to maintain close contact in providing kangaroo care after birth. Though low birth weight, prematurity, being resuscitated at birth and delivery via LSCS were found to be significantly associated with neonatal hypothermia on bivariate analysis, they were not significant in multivariable analysis. This contradicts the findings of several previous studies done in healthcare facilities in African, American and Middle Eastern countries where these risk factors were found to be significant (2–5,16–19,29,31,33). The discrepancy in these findings is likely due to the heterogeneity of the study population, study methods and settings.

Considering environmental risk factors, the monthly changes in the percentage of neonates with hypothermia tallied the temperature changes experienced by the Kandy district. Admission hypothermia was highest during the colder climate from November to January and it was lowest during the hotter climate from March to April (34). Previous studies in hospitals in Ethiopia and Nigeria have found 1.7- and 1.8 times odds of developing neonatal hypothermia in admissions during the cold season, respectively (3,15). However, the association of neonatal hypothermia with the month of delivery was not significant in our study which could have been likely due to a Type II error in research. Meanwhile, institutional factors could not be investigated in our study due to the retrospective collection of secondary data. Among them, early bathing is not practiced in the Sri Lankan setup, either in the community or in hospitals. Sri Lanka has also successfully implemented the Baby Friendly Hospital Initiative since 1992 (35). Nevertheless, more updated evidence is required on the status of skin-to-skin contact and early initiation of breastfeeding in Sri Lankan hospitals and their association with neonatal hypothermia.

Our findings showed that neonates with admission hypothermia are at a significantly higher risk for multiple complications. These findings are compatible with a study done in a health facility in Iran which found a significant rise in the development of respiratory distress, neonatal jaundice and metabolic acidosis in hypothermic neonates (22). A study done in a hospital in Malawi and a meta-analysis done in East Africa also found infant respiratory distress syndrome and neonatal health problems to be significantly associated with hypothermia respectively (5,12). In persistent hypothermia, high oxygen consumption by brown adipose tissue for non-shivering thermogenesis results in lactic acid production. This lactic acid is poorly metabolised by the immature liver which results in metabolic acidosis. Neonates are already at risk of developing jaundice due to poorly developed metabolic functions. Hypothermic neonates are at an even greater risk for jaundice due to a hypothermia-induced reduction in metabolic rate which hinders bilirubin metabolism. Furthermore, hypothermia causes high oxygen consumption, reduced surfactant release and pulmonary vasoconstriction, leading to increased respiratory distress. This also results in a higher chance of requiring ventilatory support such as CPAP (continuous positive airway pressure) ventilation (9). In our study, neonates with hypothermia showed 5.2 times the odds of mortality compared to normothermic neonates. This finding is consistent with the regional literature. Two studies done in hospitals in Iran assessing hypothermic neonates at birth showed 3.1- and 3.6 times higher odds of mortality (22,29).

This study generates recommendations for both practice and future research. For practitioners, strengthening antenatal counselling on neonatal hypothermia and its adverse effects for expectant parents with a special focus on teenage mothers, mothers with multiple pregnancies and mothers with hypertension in pregnancy is recommended. Special attention to maintaining the warm chain among day-one neonates who are being admitted is proposed as standard operating procedures for neonatal intensive care units. Raising awareness among healthcare staff on the serious morbidity and mortality of neonatal hypothermia highlighted by these findings can be recommended to promote thermo-protective interventions. Since we have linked hypothermia with both specific associated factors and adverse outcomes, future research could focus on studying the mediation effect of hypothermia in manifesting the said outcomes from possible risk factors. Since this study failed to ascertain the relationship between specific institutional and environmental factors and admission hypothermia, future studies in the South Asian region could investigate those, preferably with prospective study designs.

## Data Availability

All data produced in the present study are available upon reasonable request to the authors.

## Author’s contributions

Savindi Kasturiarachchi (SK), Inosha Alwis (IA) and Sanath Kumara (SKu) conceptualised the study. Buwanaka Rajapaksha (BR), SK and IA managed the data collection. IA conducted the data analysis. SK wrote the first draft of the manuscript, and it was read and agreed upon by all the authors.

## Data sharing statement

The datasets generated in this study can be shared upon reasonable request from the authors.

